# Linking community water and sanitation access to the global burden of antibiotic resistance using human gut metagenomes from 26 countries

**DOI:** 10.1101/2022.07.01.22277059

**Authors:** Erica R. Fuhrmeister, Abigail P. Harvey, Maya L. Nadimpalli, Karin Gallandat, Argaw Ambelu, Benjamin F. Arnold, Joe Brown, Oliver Cumming, Ashlee M. Earl, Gagandeep Kang, Samuel Kariuki, Karen Levy, Chris Pinto, Jenna M. Swarthout, Gabriel Trueba, Pablo Tsukayama, Colin J. Worby, Amy J. Pickering

**Affiliations:** Department of Civil and Environmental Engineering, Tufts University, Medford, MA, USA; Department of Civil and Environmental Engineering, University of California, Berkeley, CA, USA; Stuart B. Levy Center for Integrated Management of Antimicrobial Resistance, Tufts University, Boston, MA, USA; Department of Disease Control, London School of Hygiene and Tropical Medicine, London, United Kingdom; Faculty of Public Health, Jimma University, Jimma, Ethiopia; Francis I. Proctor Foundation, University of California, San Francisco, CA, USA; Department of Environmental Sciences and Engineering, Gillings School of Global Public Health, University of North Carolina, Chapel Hill, NC, USA; Infectious Disease & Microbiome Program, Broad Institute, Cambridge, MA, USA; The Wellcome Trust Research Laboratory, Division of Gastrointestinal Sciences, Christian Medical College, Vellore, India; Centre for Microbiology Research, Kenya Medical Research Institute, Nairobi, Kenya; Department of Environmental and Occupational Health Sciences, School of Public Health, University of Washington, Seattle, Washington, USA; Department of Global Health and Development, Faculty of Public Health and Policy, London School of Hygiene and Tropical Medicine, London, United Kingdom; Institutito de Microbiología, Colegio de Ciencias Biológicas y Ambientales, Universidad San Francisco de Quito, Quito, Ecuador; Laboratorio de Genómica Microbiana, Facultad de Ciencias y Filosofía, Universidad Peruana Cayetano Heredia, Lima, Peru; Blum Center for Developing Economies, University of California, Berkeley, CA, USA

## Abstract

**Background:** Antibiotic resistance is a leading cause of death, with the highest burden in low-resource settings. There is limited evidence on the potential for water, sanitation, and hygiene (WASH) infrastructure to reduce the burden of antibiotic resistance in humans.

**Methods:** We used geospatially tagged human gut metagenomes and household survey datasets to determine the association between antibiotic resistance gene (ARG) abundance and community-level coverage of improved drinking water points and improved sanitation facilities. Adjusted general linearized models with robust standard errors were used to estimate the relationship between ARG abundance in the human gut and access to water and sanitation.

**Findings:** We identified 1589 publicly available metagenomes from 26 countries. The average abundance of ARGs, in units of log_10_ ARG reads per kilobase per million (RPKM) mapped reads classified as bacteria, was highest in Africa compared to other World Health Organization (WHO) regions (one-way ANOVA p<0.001, post hoc Tukey HSD p<0.05). Increased access to both improved water and sanitation was associated with lower ARG abundance (effect estimate: -0.26, 95% CI [-0.44, -0.08]); the association was stronger in urban (−0.37 [-0.68, -0.07]) compared to rural areas (–0.16 [-0.38, 0.07]). Improved sanitation alone was associated with reduced ARG abundance (−0.16 [-0.32, 0.00]) while improved drinking water was not (−0.09 [-0.35, 0.16]).

**Interpretation:** While additional studies to investigate casual effects are needed, increasing access to water and sanitation could be an effective strategy to curb the proliferation of antibiotic resistance in low- and middle-income countries.

**Funding:** Bill & Melinda Gates Foundation

**Research in Context:** *Evidence before this study:* Antibiotic resistance is a growing global health threat that disproportionately affects low- and middle-income countries (LMICs). In 2019, an estimated 5 million deaths were associated with antibiotic resistance, with the highest death rate in western sub-Saharan Africa. Water, sanitation, and hygiene (WASH) interventions (e.g., household drinking water treatment, flush toilet, hand washing facilities with soap) can reduce diarrheal and respiratory infections, as reported in previous meta-analyses. Estimates, based on probability modeling, suggest improvements in water and sanitation could decrease antibiotic use for diarrheal disease treatment by 47-50% and 69-72%, respectively. Improving WASH infrastructure could theoretically contribute to the control of antibiotic resistance by preventing the release of antibiotics, resistant organisms, or antibiotic resistance genes (ARGs) into the environment, thus decreasing the burden of antibiotic-resistant infections. One global analysis across 73 countries suggested that improved infrastructure, including WASH services, was associated with reduced antibiotic resistance prevalence in isolates, however the independent effect of WASH access was not assessed. We searched PubMed for evidence on the impact of WASH interventions (excluding those related to animals and agriculture) on antibiotic resistance using the following keyword chain: *(water OR sanitation OR hygiene OR WASH) AND (antimicrobial OR antibiotic) AND resistance) NOT (“OneHealth” OR “One Health” OR animal OR livestock)*. We selected reviews and systematic reviews (n=1420) to be screened for relevance to WASH and antibiotic resistance. The reference lists of included reviews were then searched for individual studies. We also consulted international agency guidelines and online resources from the Joint Programming Initiative on Antimicrobial Resistance, the International Scientific Forum on Hygiene, ReAct, Resistomap, and the London School of Hygiene and Tropical Medicine AMR Centre. Studies focusing on centralized water or wastewater treatment technologies in high income countries (HICs) reported variable removal of antibiotics (53 to >90%), antibiotic-resistant bacteria (90-99.9%) and ARGs (90-99.9%) from waste streams. Other studies were conducted on hand hygiene, which has proven effective at reducing human infections and antibiotic use. No studies were identified on the effect of on-site sanitation systems (e.g., pour-flush toilets, pit latrines), which serve an estimated 2.7 billion people globally, or fecal sludge management interventions on antibiotic resistance.

*Added value of this study:* In this study, we used 1589 publicly available human gut metagenomes from around the world to assess the abundance of ARGs as a function of access to improved drinking water and sanitation infrastructure. This analysis provides new evidence of differences in the abundance of antibiotic resistance in the human gut across the world and finds that decreased gut abundance of ARGs is associated with increased access to improved drinking water and sanitation.

*Implications of all the available evidence:* Current approaches to controlling antibiotic resistance in humans predominantly focus on antibiotic stewardship; however, this approach is challenging in LMICs where infectious illnesses are generally more prevalent and unregulated antibiotic usage is common. Along with efforts to provide other known social benefits, such as reducing infectious disease and improving gender equality, improving access to safe drinking water and sanitation could contribute to reducing the burden of antibiotic resistance. This work highlights improving access to adequate water and sanitation as a potentially effective strategy, although additional studies designed to rigorously investigate the casual relationship between WASH and antibiotic resistance are needed.

## Introduction

Increasing rates of antibiotic resistance among bacterial pathogens are a major public health concern^1^. Many of the most concerning resistance phenotypes currently observed in the clinical setting are conferred through antibiotic resistance genes (ARGs), which can be acquired from pathogenic or commensal bacteria.^2^ In the past decade, our increased capacity to cost-effectively sequence the DNA of microbial communities (*i.e*., metagenomic sequencing) has revealed how common ARGs are among bacteria that colonize humans, animals, and environmental niches,^3^ highlighting the scale of the challenge of preventing pathogens from acquiring ARGs.

Low-and middle-income countries (LMICs) have the highest burden of antibiotic-resistant infections,^1^ and some of the most concerning ARGs (e.g., *bla*_NDM-1_ conferring beta-lactam and *mcr-1* conferring colistin resistance) are believed to have first mobilized into human pathogens in LMICs.^4,5^ Inadequate water, sanitation and hygiene (WASH) infrastructure could exacerbate the spread of resistance.^6^ Clean drinking water and flush toilets are nearly universal in most high-income countries (HICs), but variable to non-existent in many LMIC settings. Access to improved WASH could prevent the emergence, transfer and spread of antibiotic resistance in many ways.^7^ Here, we have depicted potential interventions to stop spill over between humans, animals, the environment, and food supply systems (**Figure 1**). First, use of toilets (improved sanitation) and wastewater treatment greatly limits the load of antibiotic-resistant bacteria and ARGs that could otherwise be excreted into the environment. *E. coli*, which is enriched in excreta was implicated in 23% of total global deaths attributable to antibiotic resistance in 2019.^1^ Uninterrupted access to clean drinking water provides an additional barrier to exposure. Second, by reducing the overall infectious disease burden and accompanying need for antibiotic use, WASH services could indirectly reduce the population-level antibiotic selection mechanisms that support the propagation of resistant strains.^8^ Previous global analyses leveraging urban sewage samples have found that country-level WASH access is associated with ARG burden, lending support to this hypothesis.^9^ However, given that the majority of human fecal waste generated in LMICs does not enter centralized sewer systems,^10^ investigations that rely on urban sewage samples are not representative.

**Figure 1.**
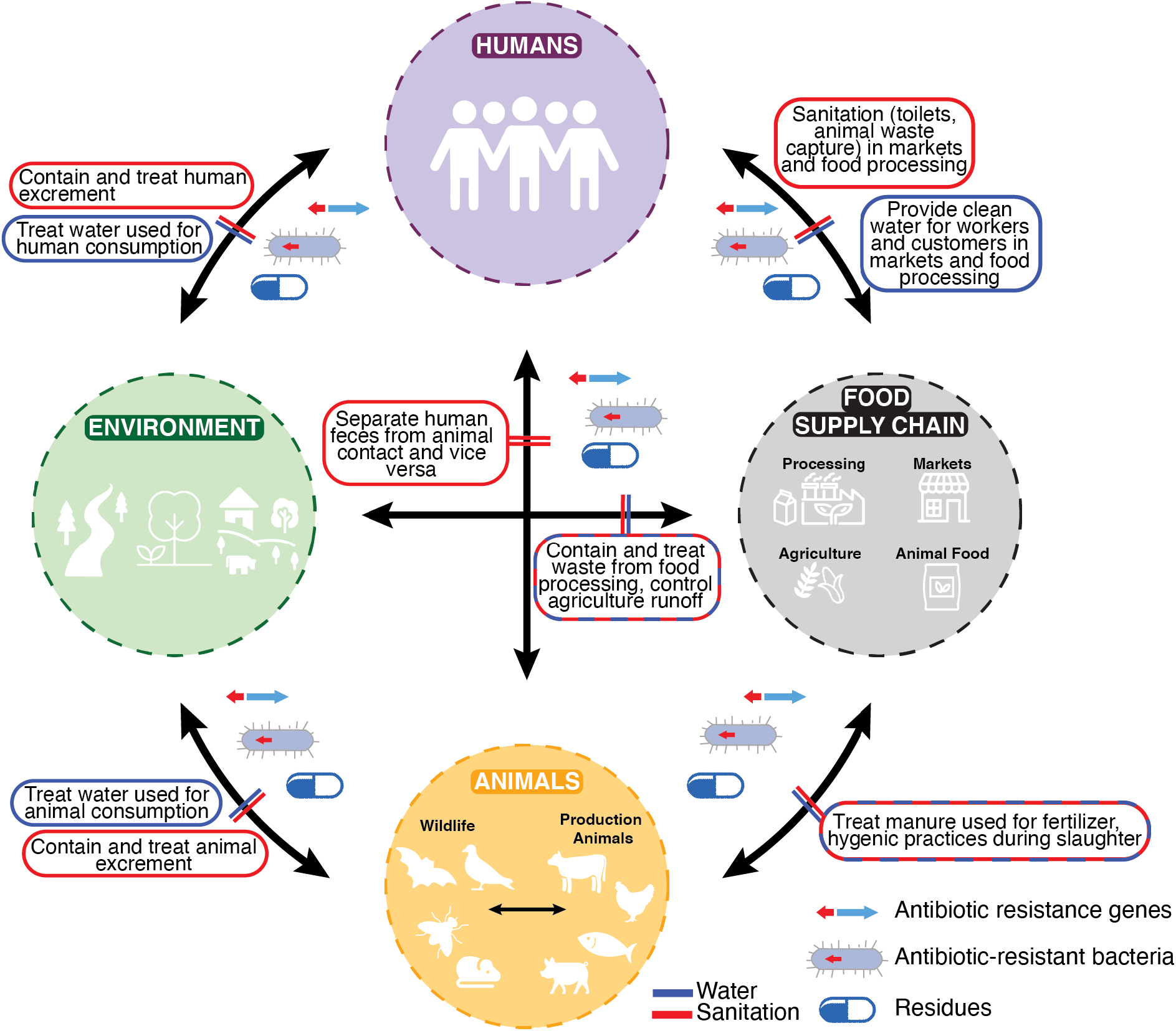
Potential water (blue) and sanitation (red) interventions needed to block the transmission of antibiotic resistance genes and antibiotic-resistant bacteria as well as dissemination of antibiotic residues between humans, animals, the environment, and the food supply chain.

Here, we leverage publicly available human gut metagenomes worldwide to describe regional trends and investigate the association between community levels of drinking water and sanitation access and ARG abundance in the human gut. Gut metagenome data from global populations have become increasingly available over the past decade, including from LMICs that lack centralized water and sanitation infrastructure. The overall goal of our analysis was to examine regional differences in ARG abundance and contribute new evidence on whether limited WASH infrastructure may be a critical driver of human antibiotic resistance carriage.

## Methods

### Metagenome Identification

We obtained publicly available human gut metagenomes, resulting from short-read (Illumina) whole-genome sequencing, from the NCBI sequence read archives (SRA). We included all available metagenomes from LMICs and a subset of metagenomes from HICs. Studies from HICs were selected to span varying human ages and geographic locations. Longitude and latitude coordinates were obtained from the SRA metagenome metadata and verified with the corresponding research article. If coordinates were unavailable in the SRA, location was determined based on the description in the research article. Additional human gut metagenomes were contributed from Mozambique, Bangladesh, and Kenya by authors of these studies. Metagenomes were excluded if location and combined access to both drinking water and sanitation could not be determined.

### Antibiotic Resistance Gene Identification and Normalization

ARGs were identified by mapping reads to the Comprehensive Antibiotic Resistance Database (CARD)(v 3.0.9)^11^ of ARG protein sequences using BLASTX in DIAMOND (v 0.9.30.131).^12^ Results were filtered using an alignment cutoff of 25 amino acids and an identity of 95%. Paired-end reads were mapped separately, and duplicate mappings were removed. To normalize for total bacteria in each sample, reads per kilobase per million mapped reads (RPKM) of ARGs were calculated using the total number of reads classified as bacteria (using Bracken^13^) for total reads.

### Taxonomy

Kraken2^14^ with the standard database was used for taxonomic classification of short reads. Bracken^13^ corrected counts were used to calculate the total number of reads classified as bacteria as well as the relative abundance of the five most abundant families among reads classified as bacteria.

### Survey Methods

Household variables, including access to improved drinking water and sanitation, were primarily obtained from geospatially tagged, nationally representative household survey data sets. Demographic and Health Surveys (DHS) (https://www.dhsprogram.com) were obtained for the year closest to the study date (2010-2018). When DHS data were not available (El Salvador, Mongolia, and Mexico), we obtained Multiple Indicator Cluster Surveys (MICS) (http://mics.unicef.org/surveys) from the closest available year to the study date (appendix Table A1). For the countries without DHS or MICS data (Ecuador, China), we obtained comparable country-level or study-specific survey data (Table A1). If the study year was unknown (N=9), we used the most recent survey available.

For countries with DHS data (which includes GPS coordinates for survey clusters), survey clusters located within a 25 km radius of the metagenome coordinates were selected. Two additional radii (50 km and 75 km) were used to conduct a sensitivity analysis of radius threshold. For countries without DHS data (which contains information on the region or district), we determined the locally defined administrative area containing the metagenome coordinate and filtered to households within that administrative area. Within the defined geographic region (radius or administrative area), we selected urban, rural, or both types of household data based on the authors’ description of the study site. From survey data joined to each metagenome dataset, we calculated the proportion of households in the geographically defined radius with access to improved sanitation (separates excreta from human contact), access to improved drinking water (design protects source from contamination), and access to both improved sanitation and improved drinking water (definitions in appendix Table A2). We were unable to investigate access to safely managed sanitation and drinking water, as defined by the Sustainable Development Goals,^15^ due to the absence of estimates for most LMICs in our analysis. Since access to improved drinking water and sanitation could not be independently determined for HICs, we assigned a value of 99% for combined access. We calculated the proportion of children using antibiotics based on reported use in response to diarrhea or fever in the previous two weeks. In addition, we determined the proportion of households owning livestock and other assets. We used consumption estimates (Defined Daily Dose (DDDs) per 1000 people per day) to estimate country-level antibiotic usage. Additional data obtained included antibiotic consumption estimates (DDDs per 1000 people per day)^16^, gross domestic product (GDP) per capita^17^, income classification per country as determined by World Bank designation in 2021, population density^18^, and animal antibiotic consumption in 2010.^19^

### Data Analysis

We used generalized linear models to model log_10_-transformed, normalized abundance of ARGs (in total, by drug class, and individually) as a function of local access to both improved sanitation and improved drinking water in R (v 4.0.5). In separate analyses, total abundance was modeled as a function of local access to improved drinking water and improved sanitation individually. Robust standard errors were used to account for clustered data (multiple samples in one georeferenced cluster). World Health Organization (WHO) region and population density were included as covariates in all models. Two adjusted analyses were conducted. First, we included additional covariates that were available for the full set of metagenomes (antibiotic usage in humans, GDP per capita, library layout-paired or single-, and average read length). A second adjusted analysis was conducted for the subset of samples with additional data available (shared sanitation, walk time to water source, antibiotic consumption in children, animal antibiotic consumption, livestock ownership-any animal-, age, sex, finished walls, floors, roof, electricity access, watch/clock, radio, TV, mobile phone, refrigerator, bike, motorcycle, car/truck, and clean fuel). The association between each covariate and outcome variable was assessed; those with a p-value < 0.2 were screened for collinearity. A subset of prescreened variables was included in the final model to reduce collinearity. Our prespecified analysis plan is available on Open Science Framework (https://osf.io/n4z7f/).

Associations between abundance of ARGs and access to improved water and sanitation were conducted in subsets of the data separated by age (0-18 vs.18+), World Bank income classification (low, lower-middle vs. upper-middle, high), WHO region, animal antibiotic usage (< 50 vs. ≥ 50 mg/population-corrected unit, PCU), urbanicity (rural vs. urban), relative abundance of *Enterobacteriaceae* (< 2% vs. ≥ 2%), and human antibiotic usage (< 12 vs. ≥ 12 DDD per 1000 persons per day).

For models of individual gene abundance, genes present in less than 5% of samples were excluded and p-values were corrected for multiple comparisons using the Bonferroni method.

We used one-way ANOVA followed by post hoc Tukey HSD to compare abundance of ARGs and log_10_-transformed relative abundance of *Enterobacteriaceae* by region. To compare total ARGs by urbanicity in each WHO region, we used student t-tests and a Bonferroni correction.

## Results

### Identified Study Characteristics

We identified 36 studies in total: 25 studies in the SRA containing metagenomes from LMICs, three with metagenomes from LMICs from authors of this work, and eight from HICs. Three studies (2 China, 1 Fiji) were excluded due to lack of data on both improved drinking water and sanitation. One study (Brazil) was omitted due to a high relative abundance of non-bacterial taxa and no detectable ARGs in several metagenomes. In total, 1589 metagenomes from 26 countries (4 low, 8 lower-middle, 6 upper-middle, and 8 HIC) were included in the analysis (**Figure 2A**, and appendix Table A3).

**Figure 2.**
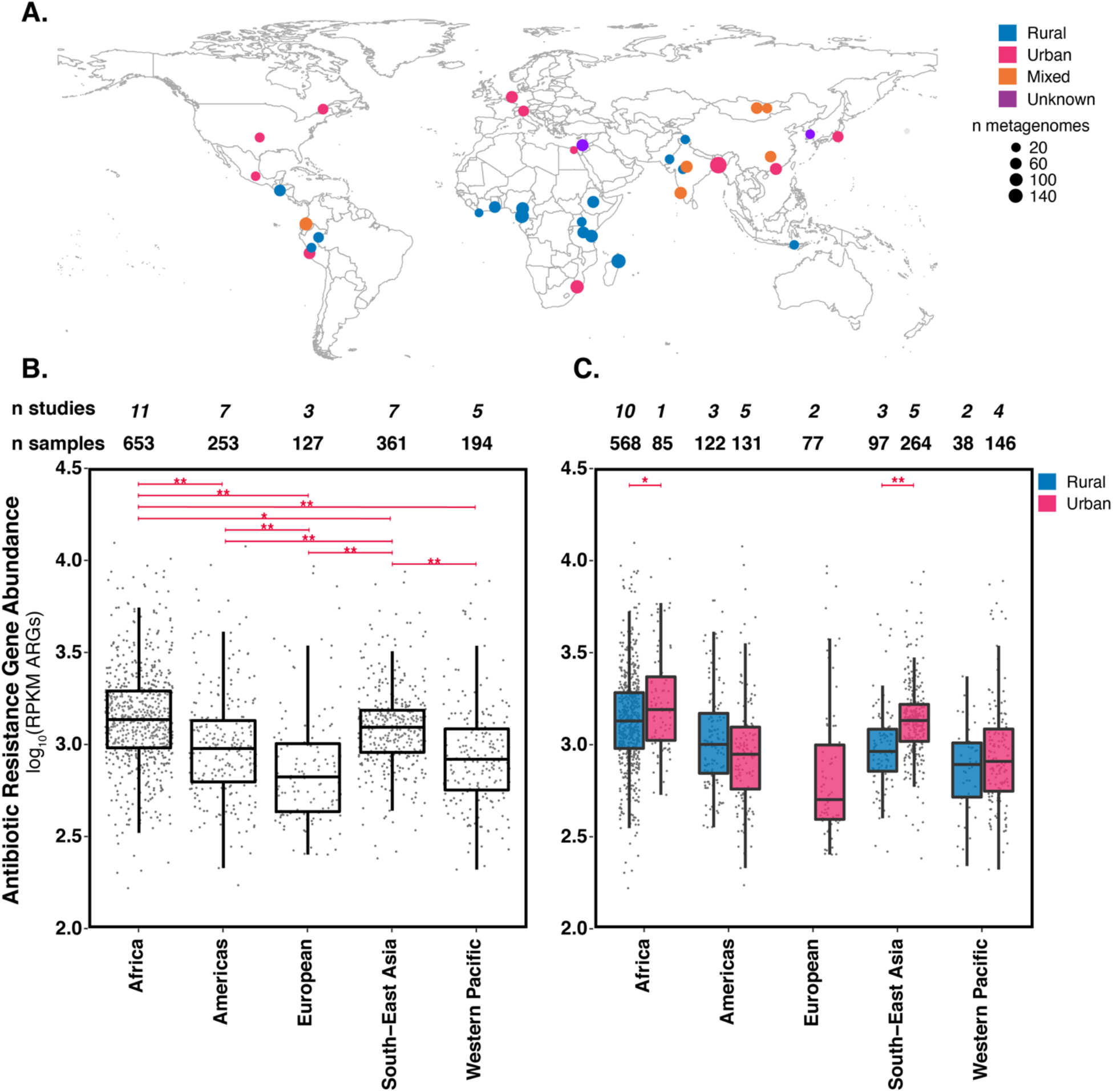
A) Location of studies included in this analysis. Urbanicity indicated by color and number of metagenomes indicated by size of dot. Additional study information in appendix Table A3. B) Abundance of antibiotic resistance genes in units of log10ARG reads per kilobase per million (RPKM) mapped reads classified as bacteria by WHO region. **Indicates *p*<0.001 and * indicates p<0.05 by ANOVA with post hoc Tukey HSD. C) Abundance of antibiotic resistance genes in rural and urban areas by WHO region. **Indicates *p*<0.001 and * indicates p<0.05 by student’s t-test correcting for multiple comparisons. Boxplots consist of 25^th^ percentile, median, 75^th^ percentile and whiskers extend to at most 1.5x the inner quartile range. Eastern Mediterranean is not shown in B and C due to the presence of only 1 metagenome (Egypt). Rural European is not shown in C, as no metagenomes were identified from rural Europe.

The most abundant bacterial families among all metagenomes were, *Prevotellaceae, Bacteroidaceae, Oscillospiraceae* (commonly referred to as *Ruminococcaceae*), *Lachnospiraceae*, and *Enterobacteriaceae* (Figure A1). On average, the most abundant family in Africa and South-East Asia (SEA) was *Prevotellaceae* whereas Europe and the Western Pacific were dominated by *Bacteroidaceae*. There was a high degree of variability in metagenomes in the Americas with *Prevotellaceae* most abundant in El Salvador and Ecuador, *Bacteroidaceae* most abundant in Canada, and *Oscillospiraceae* most abundant in Peru. Since *Enterobacteriaceae* are known to harbor a disproportionately high number of ARGs and ARG databases are dominated by *Enterobacteriaceae* alleles, we were interested in the variation in relative abundance across WHO regions. The log_10_-transformed average relative abundance of *Enterobacteriaceae*, ranging from -0.03 ± 0.7 [mean ± std] in Europe to 0.5 ±0.6 in Africa, was higher in Africa compared to all other WHO regions (one-way ANOVA p<0.001; post hoc Tukey HSD p<0.05 for SEA and p<0.001 for Americas, Europe, and Western Pacific). Notably, the average relative abundance of *Enterobacteriaceae* was above 15% in Mozambique, Madagascar, Italy, Brazil, Indonesia, and in one Bangladesh study.

### Abundance of Antibiotic Resistance Genes

The average log_10_-transformed, normalized abundance of ARGs was highest in Africa (mean=3.14) compared to all other WHO regions (one-way ANOVA p<0.001, post hoc Tukey HSD p<0.001 for all but SEA, p=0.03 for SEA; **Figure 2B**), followed by SEA which was significantly higher than the Americas, Europe, and Western Pacific (p<0.001). Abundance trends varied by drug class (appendix Figure A2). Beta-lactamase genes were highest in Africa and SEA (appendix Figure A2). Tetracycline resistance was ubiquitous throughout all WHO regions (**Figure 3** and appendix Figure A3). Beyond genes conferring resistance to tetracyclines, the resistome (collection of all ARGs in a metagenome) was dominated by genes encoding resistance to beta-lactams in Africa, Europe, and some countries in the Americas. In SEA resistance to beta-lactams and macrolides, lincosamides, and streptogramins (MLS) were dominant, while MLS were abundant in the Western Pacific. Among metagenomes where urbanicity could be determined, there was a higher ARG abundance in urban areas in Africa and SEA (SEA p<0.001, Africa p=0.01; **Figure 1C**).

**Figure 3.**
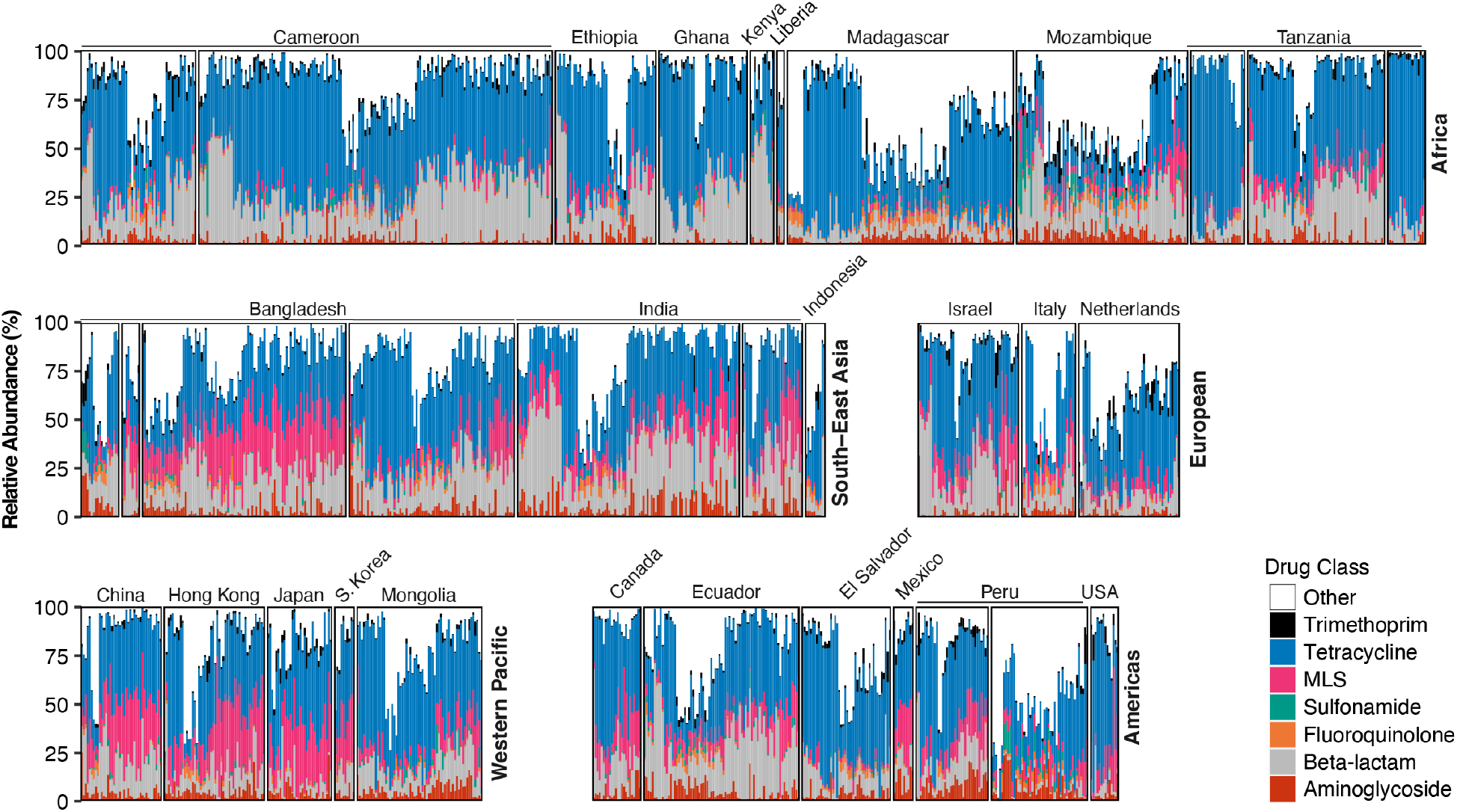
Relative abundance of antibiotic resistance genes by drug class. Metagenomes ordered by hierarchal clustering of relative abundance by drug class profiles and grouped by study as well as WHO region. Each bar represents the relative proportion of each drug class in one metagenome. Eastern Mediterranean is not shown due to the presence of only 1 metagenome (Egypt). Note: multiple studies conducted in Cameroon, Tanzania, Bangladesh, India, and Peru. MLS=Macrolide, lincosamide, streptogramin.

### Association between ARG Abundance and Access to Improved Water and Sanitation

Overall, increased access to improved water and sanitation was associated with a decrease in ARG abundance (**Table 1**). A 100% increase in access to improved water and sanitation was associated with a 0.26 reduction in ARG abundance (−0.26 [-0.44, -0.08], p<0.01) (estimate [95% CI]) in the analysis adjusting for all covariates. A more realistic increase in access to water and sanitation of 25% was associated with a 0.07 reduction in ARG abundance. Relaxing the geographical linkage between survey data and reported sample location (using 50 and 75km radii) did not significantly impact results; nor did the incorporation of additional covariates where available in the adjusted analysis (appendix Tables A4 and A5). Across quartiles of water and sanitation access, significantly lower ARG abundance was found in regions with 75-100% access compared to 0-25% access (−0.17 [-0.34, 0.00], p=0.04).

**Table 1.**
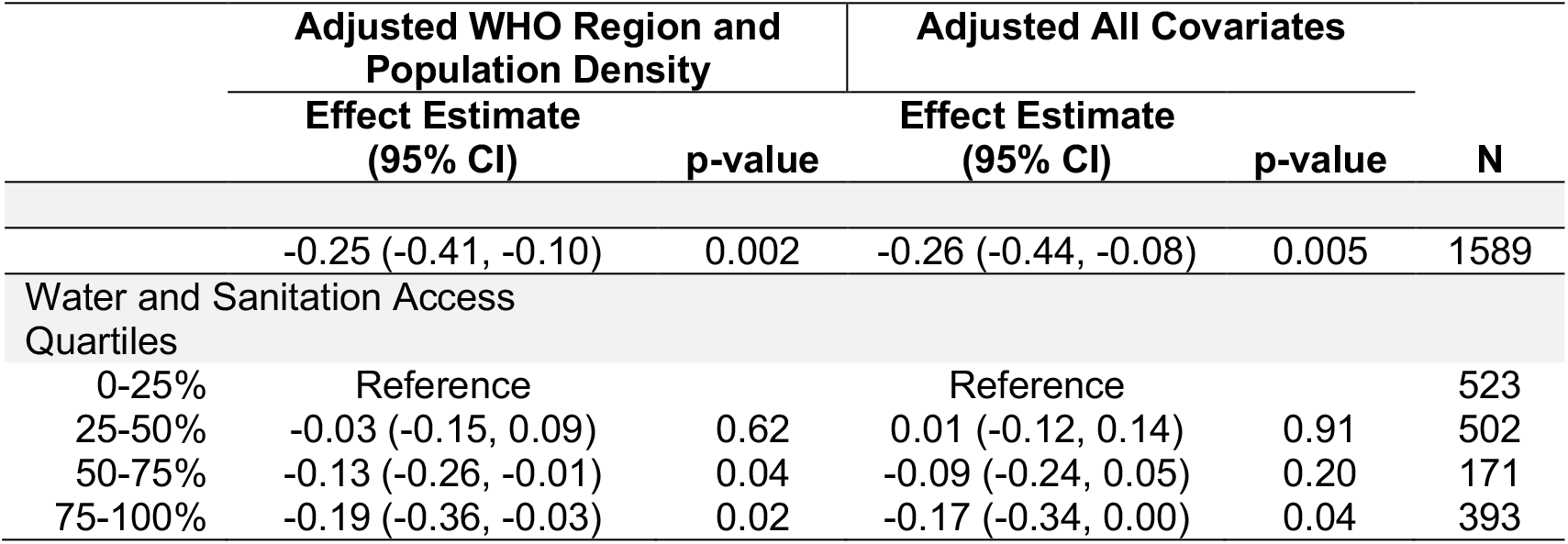
Association between ARG abundance and combined access to both improved drinking water and sanitation. Adjusted analyses were conducted with WHO region and population density as covariates (Adjusted WHO Region and Population Density) and additional covariates of GDP per capita, antibiotic usage in humans, read length, and library layout (Adjusted All Covariates). Additional information on covariates provided in appendix Table A2. Effect estimates were generated across the full range and by quartiles of improved water and sanitation coverage. Robust standard errors were used to account for clustering (multiple metagenomes at one georeferenced cluster).

The magnitude of association was higher for improved sanitation access alone compared to improved drinking water alone (sanitation: -0.16 [-0.32, 0.00] p=0.05; drinking water: -0.09 [-0.35, 0.16], p=0.47; appendix Table A6).

Considering the most common drug classes (those detected in ≥80% of samples), we found increased access to improved water and sanitation was associated with a lower abundance of ARGs conferring resistance to tetracycline (−0.31 [-0.48, -0.13], p<0.001), fluoroquinolones (−0.74 [-1.40, -0.08], p=0.03) and trimethoprim (−1.00 [-1.70, -0.31], p<0.01) antibiotics (appendix Table A7), while WASH was not associated with abundance of genes conferring resistance to beta-lactams, aminoglycosides, or MLS (p>0.05 for all).

Improved water and sanitation was associated with a greater decrease in ARG abundance in urban compared to rural areas (urban: -0.37 [-0.68, -0.07]; rural: -0.16 [-0.38, 0.07], p_interaction_=0.26 **Table A8)**. There was a greater, but non-significant, decrease, in higher-income compared to lower-income countries, areas with higher antibiotic usage in animals, and areas with higher antibiotic usage in humans. In the subset of metagenomes with higher relative abundance of *Enterobacteriaceae*, there was a greater decrease in ARG abundance compared to metagenomes with a lower relative abundance (RA) (RA ≥ 2%: -0.29 [-0.45, -0.12]; RA < 2%: - 0.18 [-0.33, -0.03], p_interaction_=0.80). Of the WHO regions investigated, improved water and sanitation was associated with the greatest decrease in ARGs in the Americas (−0.56 [-0.91, - 0.21], p<0.001) and SEA (−0.28 [-0.52, -0.03], p=0.03).

Increased access to improved water and sanitation was associated with a decrease in the abundance of 29 resistance genes conferring resistance to a range of antibiotic classes in the analysis adjusted for all covariates (**Figure 4** and appendix Table A9). Of those 29 ARGs, *dfrA1* (conferring resistance to trimethoprim), *tetL* and *tetM* (tetracycline), *ermT* (MLS), *qnrS* (fluoroquinolone), and *bla*_CTX-M_ Group 2 (beta-lactamases) have been identified as current health threats due to their presence in human-associated environments, carriage by mobile genetic elements, and presence in the genomes of ESKAPE (*Enterococcus faecium, Staphylococcus aureus, Klebsiella pneumoniae, Acinobacter baumannii, Pseudomonas aeruginosa*, and *Enterobacter*) pathogens.^20^

**Figure 4.**
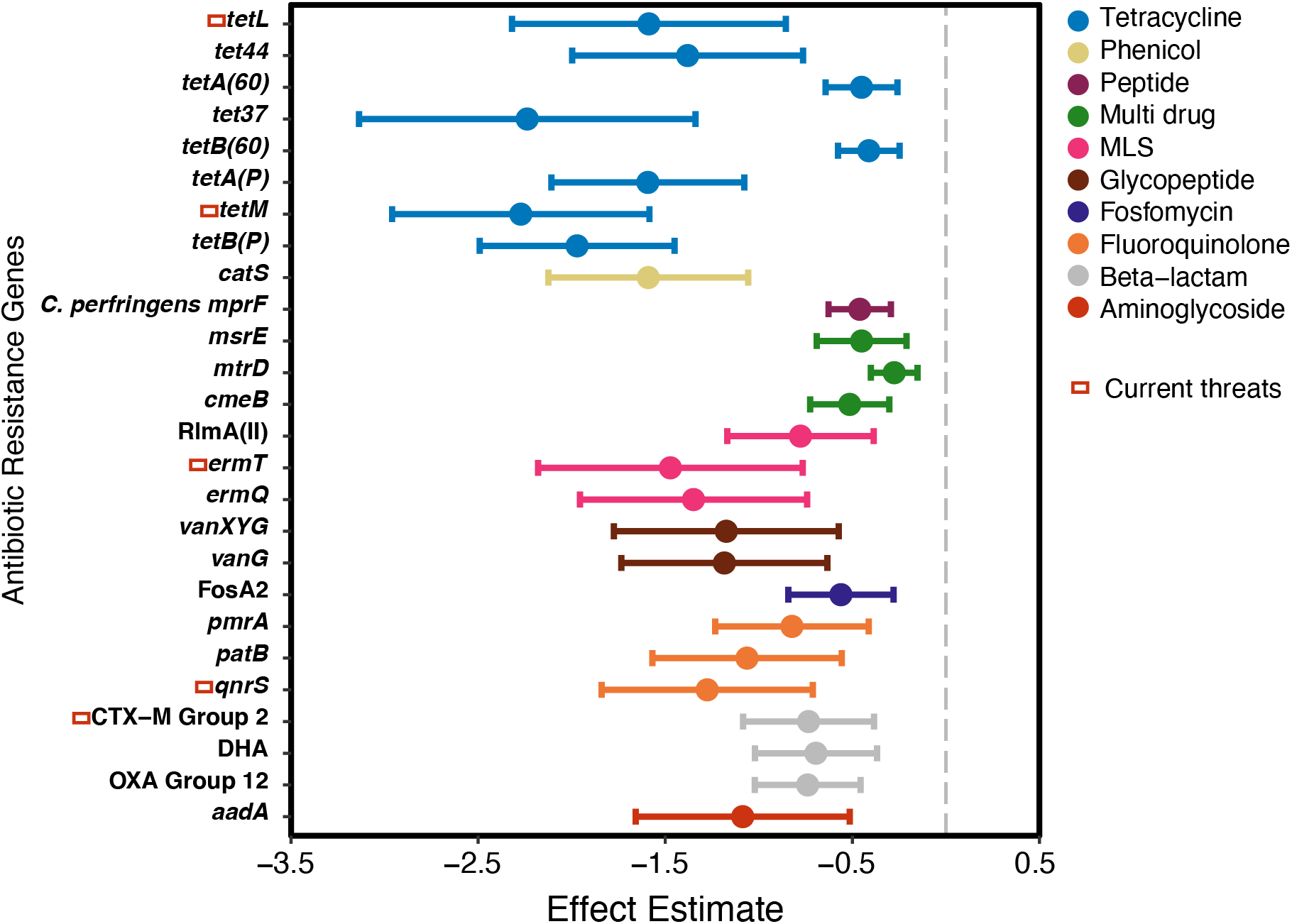
Association between abundance of individual ARGs and combined access to both improved drinking water and sanitation. Effect estimate (change in log10 ARG abundance for a 100% increase in access to improved water and sanitation) and 95% confidence intervals for antibiotic resistance genes present in at least 5% of metagenomes and significant at p-value <0.05 in the fully adjusted model. Robust standard errors were used to account for clustering (multiple metagenomes at one georeferenced cluster). P-values were corrected for multiple comparisons using the Bonferroni method. Zero values were assigned a value of half the lowest gene abundance. ARGs that are enriched in human-associated environments, mobile, and present in ESKAPE pathogens are referred to as “Current threats”^20^ and are indicated by the leftmost rectangle. Beta-lactam resistance genes included in each group are available in appendix Table A9. MLS=Macrolide, lincosamide, streptogramin.

## Discussion

We found that increased access to improved water and sanitation was associated with decreased ARG abundance, particularly in urban areas, and areas with higher antibiotic usage in humans and animals. We were unable to identify any metagenomes explicitly from rural sites in HICs, making it difficult to isolate the impact of urbanicity from economy type. Improved sanitation was associated with a greater decrease in ARG abundance compared to drinking water alone, but the greatest decrease was observed with access to both improved drinking water and sanitation. Our results suggest increasing access to improved water and sanitation at the community-level could be an effective strategy to reduce the proliferation of antibiotic resistance. Additional studies are needed to determine if there is a causal relationship between improving WASH and antibiotic resistance burden. Also, we identified a relatively small number of human gut metagenomes from around the world, especially in LMICs. There is a clear need for more metagenomic data from LMICs where the burden of infectious illnesses is highest and microbiome-based research has the potential to improve human health.

The abundance of ARGs was similar in magnitude in all WHO regions, but abundance was highest in Africa. All studies in the WHO-defined African region were from sub-Saharan Africa, and thus abundance estimates are not representative of North Africa which benefits from better WASH conditions. Total ARGs were also highest in Africa using wastewater surveillance in urban areas,^9^ which is notable given most metagenomes included in our analysis from Africa were from rural settings. We found the lowest abundance of ARGs in the Americas. Unlike Africa, the Americas were relatively heterogeneous in terms of most abundant taxa (including *Enterobacteriaceae*) and classes of ARGs. Previous urban wastewater surveillance efforts found the Americas had the second highest ARG abundance,^9^ but significant variation in gut microbiomes by region and urbanicity in the Americas could contribute to discrepancies in resistome characterization between our work and others. Tetracycline resistance was abundant in all WHO regions, consistent with a previous analysis of human gut metagenomes from 11 countries.^21^ The widespread distribution of tetracycline resistance is not surprising given it has been used globally for decades as a broad-spectrum antibiotic in humans and animals.^22^ Lastly, there was a relatively high abundance of genes conferring resistance to beta-lactams in Africa and South-East Asia. Traveler studies indicate that northern Africa and Asia are reservoirs of beta-lactam resistance, including extended-spectrum β-lactamase-producing *Enterobacteriaceae*.^23^

Our findings, in line with previous literature, suggest increasing access to improve water and sanitation services within communities is a potential strategy to combat antibiotic resistance. A metagenomic analysis of wastewater samples from 60 high- and low-income countries found that lower open defecation rates, higher access to improved water sources in rural areas, and increased water and sanitation investments were associated with decreased ARG abundance.^9^ We found similar associations between ARG abundance and water and sanitation access in our analysis of human gut metagenomes from rural and urban areas in LMICs, which are likely poorly represented in wastewater surveillance efforts given that 91% of rural and 37% of urban populations are not connected to sewerage systems.^15^ Modeling studies have also reported associations between antibiotic resistance and WASH. In a study of 73 countries around the world with antibiotic consumption data, infrastructure improvements - including but not limited to improved water and sanitation – were associated with reduced prevalence of antibiotic resistant organisms (*Escherichia coli, Klebsiella* spp., and *S. aureus*) in clinical surveillance data.^6^

Similarly, combined access to both improved water and sanitation resulted in the greatest reduction in ARG abundance in our analysis where we captured diversity of ARGs circulating in community rather than clinical settings. A modelling-based investigation of the potential impact of varying levels of sanitation improvements on abundance of ARGs in the environment in South-East Asia found that moving from open defecation to improved sanitation, in general, provided the greatest reduction in ARGs, with proportionally smaller reductions occurring as more advanced wastewater treatment processes were added.^24^ Lastly, in a country-specific, experimental analysis (in contrast to the aforementioned global or WHO regional-scale, observational studies) there was no association between WASH and ARG carriage, measured using PCR one year after the intervention, among 120 children (<14 months old) in Maputo, Mozambique.^25^ Additional studies across diverse populations are needed to further investigate the relationship between WASH and antibiotic resistance gene abundance in human guts. The period of follow-up in future intervention studies should also consider amount of time needed to clear pre-existing antibiotic resistance genes from the human gut.

An important limitation to our work is the observational, ecological design and the potential for confounding. While our study adjusted for WHO region in all models to account for confounding socioeconomic variables, it is possible our estimates are an overestimate of the association between ARG abundance and water and sanitation. Other limitations include a lack of standardized surveys collecting data on antibiotic consumption, both quantity and diversity, of antibiotics across communities. Current sources are constrained to commercial sales data or household reported consumption in sick children, both of which are limited in scope. Additionally, *Enterobacteriaceae* growth in the absence of temperature control or preservation solutions during sample collection and transport could impact our results, although most metagenomic studies tightly control sample collection, transport, and testing. In the subgroup analysis there was a stronger association between access to improved water and sanitation and ARG abundance in metagenomes with higher relative abundance of *Enterobacteriaceae*. Also, identification of ARGs through alignment-based methods is known to result in false positives. In a small subset of metagenomes, we used ROCker^26^, a modeling-based strategy, to identify Class A beta-lactamase genes and found 75% agreement between CARD identifications and ROCker (data not shown). While we chose to normalize our estimates of ARG abundance using total bacterial reads, this does not account for bacterial genome size variation among samples and could provide higher estimates when ARGs are carried on smaller genomes. Alternative approaches leveraging read mapping to bacterial single copy core genes (SCGs)^27^ attempt to correct for this, though may not account for confounders such as plasmid copy number variability and incomplete carriage of the chosen SCG set. To identify any potential discrepancies, we performed a secondary analysis using a SCG normalization approach and did not find the results to differ considerably although the magnitude of effect was somewhat diminished (SI pg 3). Finally, use of the CARD database could overstate abundance of *Enterobacteriaceae* and presence of antibiotic resistance genes does not imply phenotypic resistance.

Education and better regulated use of antimicrobials in humans and animals (i.e., stewardship) are the main approaches to managing antibiotic resistance. Mitigating infectious diseases is also a critical component of the WHO Global Action Plan on Antimicrobial Resistance, which recommends strategies such as vaccinations and effective WASH.^28^ However, the focus for WASH has been on healthcare rather than community settings. In a review of the 77 action plans on antimicrobial resistance in the WHO library, only 11 countries mention community-level WASH.^29^ While most surveillance on antimicrobial-resistant organisms has been limited to clinical settings in LMICs, community acquired resistance is substantial.^30^ Incorporating WASH infrastructure in community settings into national strategies could be effective for curbing antibiotic resistance in LMICs. We found the most significant reduction in ARGs was associated with access to combined improvements in water and sanitation, suggesting comprehensive WASH access may be more effective than single interventions.

## Data Availability

All raw sequencing reads are available from the Sequence Read Archives. Data used for analyses are available on open science framework:(https://osf.io/n4z7f/)

## Contributors

AJP, ERF, and MLN conceptualized the study. APH and ERF contributed to data curation. ERF conducted the analysis which was replicated by APH and JMS. AJP, JB, and KL provided additional data (metagenomes and/or metadata) beyond those publicly available at the time of analysis. CP contributed to the conceptual diagram. KG conducted the literature review. ERF, MLN, and KG contributed to the first draft of the manuscript. All co-authors provided input on methods and edited the final manuscript. AJP and OC obtained funding for the study.

## Acknowledgments

This work was supported by grant LSHTM INV-006556 from the Bill and Melinda Gates Foundation. ERF was supported by the NSF Postdoctoral Research Fellowships in Biology Program under Grant No. 1906957. MLN was supported by NIH award KL2TR002545 and the Stuart B. Levy Center for Integrated Management of Antimicrobial Resistance at Tufts (Levy CIMAR), a collaboration of Tufts Medical Center and the Tufts University Office of the Vice Provost for Research (OVPR) Research and Scholarship Strategic Plan (RSSP). KL and GT contributions were supported by NIAID grant numbers R01AI137679 and 1K01AI103544. AME and CJW’s contributions were supported by NIAID grant number U19AI110818. SK is supported by the National Institute of Allergy and Infectious Diseases of the National Institutes of Health under Award Number R01AI099525 and the Wellcome Trust Grant Number 215675-Z-19-Z. The authors would also like to thank Konstantinos Konstantinidis for his consultation on bioinformatic methods. Any opinions, findings, and conclusions or recommendations expressed in this material are those of the author(s) and do not necessarily reflect the views of the funding organizations.

## Data Sharing

SRA project numbers for studies used in this analysis are located in Table A3. Compiled data used in our analysis are available on OSF.

## References

1 Murray CJ, Ikuta KS, Sharara F, et al. Global burden of bacterial antimicrobial resistance in 2019: a systematic analysis. The Lancet 2022; 399: 629–55.

2 U.S. Department of Health and Human Services, Centers for Disease Control and Prevention (CDC). Antibiotic Resistance Threats in the United States, 2019. Atlanta, GA, 2019.

3 Pal C, Bengtsson-Palme J, Kristiansson E, Larsson DGJ. The Structure and Diversity of Human, Animal and Environmental Resistomes. Microbiome 2016; 4: 54.

4 Kumarasamy KK, Toleman MA, Walsh TR, et al. Emergence of a new antibiotic resistance mechanism in India, Pakistan, and the UK: a molecular, biological, and epidemiological study. Lancet Infect Dis 2010; 10: 597–602.

5 Liu Y-Y, Wang Y, Walsh TR, et al. Emergence of plasmid-mediated colistin resistance mechanism MCR-1 in animals and human beings in China: a microbiological and molecular biological study. Lancet Infect Dis 2016; 16: 161–8.

6 Collignon P, Beggs JJ, Walsh TR, Gandra S, Laxminarayan R. Anthropological and socioeconomic factors contributing to global antimicrobial resistance: a univariate and multivariable analysis. Lancet Planet Health 2018; 2: e398–405.

7 Nadimpalli ML, Marks SJ, Montealegre MC, et al. Urban informal settlements as hotspots of antimicrobial resistance and the need to curb environmental transmission. Nat Microbiol 2020; 5: 787–95.

8 Lipsitch M, Samore MH. Antimicrobial Use and Antimicrobial Resistance: A Population Perspective. Emerg Infect Dis 2002; 8: 347–54.

9 Hendriksen RS, Munk P, Njage P, et al. Global monitoring of antimicrobial resistance based on metagenomics analyses of urban sewage. Nat Commun 2019; 10: 1124.

10 Jones ER, van Vliet MTH, Qadir M, Bierkens MFP. Country-level and gridded estimates of wastewater production, collection, treatment and reuse. Earth Syst Sci Data 2021; 13: 237– 54.

11 Alcock BP, Raphenya AR, Lau TTY, et al. CARD 2020: antibiotic resistome surveillance with the comprehensive antibiotic resistance database. Nucleic Acids Res 2020; 48: D517–25.

12 Buchfink B, Xie C, Huson DH. Fast and sensitive protein alignment using DIAMOND. Nat Methods 2015; 12: 59–60.

13 Lu J, Breitwieser FP, Thielen P, Salzberg SL. Bracken: estimating species abundance in metagenomics data. PeerJ Comput Sci 2017; 3: e104.

14 Wood DE, Lu J, Langmead B. Improved metagenomic analysis with Kraken 2. Genome Biol 2019; 20: 257.

15 WHO and UNICEF. Progress on drinking-water, sanitation and hygiene: 2017 update and SDG baselines. 2017 https://www.who.int/publications-detail-redirect/9789241512893 (accessed June 23, 2021).

16 Browne AJ, Chipeta MG, Haines-Woodhouse G, et al. Global antibiotic consumption and usage in humans, 2000–18: a spatial modelling study. Lancet Planet Health 2021; 5: e893– 904.

17 The World Bank. GDP (current US$) Data. https://data.worldbank.org/indicator/NY.GDP.MKTP.CD (accessed June 23, 2021).

18 Center For International Earth Science Information Network-CIESIN-Columbia University. Gridded Population of the World, Version 4 (GPWv4): Population Density, Revision 11. 2017. DOI:10.7927/H49C6VHW.

19 Antibiotic use in livestock. Our World Data. https://ourworldindata.org/grapher/antibiotic-use-in-livestock (accessed June 17, 2021).

20 Zhang A-N, Gaston JM, Dai CL, et al. An omics-based framework for assessing the health risk of antimicrobial resistance genes. Nat Commun 2021; 12: 4765.

21 Feng J, Li B, Jiang X, et al. Antibiotic resistome in a large-scale healthy human gut microbiota deciphered by metagenomic and network analyses. Environ Microbiol 2018; 20: 355–68.

22 Roberts MC. Tetracycline Therapy: Update. Clin Infect Dis 2003; 36: 462–7.

23 Ruppé E, Andremont A, Armand-Lefèvre L. Digestive tract colonization by multidrug-resistant Enterobacteriaceae in travellers: An update. Travel Med Infect Dis 2018; 21: 28–35.

24 Graham DW, Giesen MJ, Bunce JT. Strategic Approach for Prioritising Local and Regional Sanitation Interventions for Reducing Global Antibiotic Resistance. Water 2019; 11: 27.

25 Berendes D, Knee J, Sumner T, et al. Gut carriage of antimicrobial resistance genes among young children in urban Maputo, Mozambique: Associations with enteric pathogen carriage and environmental risk factors. PloS One 2019; 14: e0225464.

26 Orellana LH, Rodriguez-R LM, Konstantinidis KT. ROCker: accurate detection and quantification of target genes in short-read metagenomic data sets by modeling sliding-window bitscores. Nucleic Acids Res 2017; 45: e14.

27 Lee K, Kim D-W, Lee D-H, et al. Mobile resistome of human gut and pathogen drives anthropogenic bloom of antibiotic resistance. Microbiome 2020; 8: 2.

28 World Health Organization. Global action plan on antimicrobial resistance. 2015 https://www.who.int/publications-detail-redirect/9789241509763 (accessed Aug 26, 2021).

29 Essack S. Water, sanitation and hygiene in national action plans for antimicrobial resistance. Bull World Health Organ 2021; 99: 606–8.

30 Ingle DJ, Levine MM, Kotloff KL, Holt KE, Robins-Browne RM. Dynamics of antimicrobial resistance in intestinal Escherichia coli from children in community settings in South Asia and sub-Saharan Africa. Nat Microbiol 2018; 3: 1063–73.

