## Supplemental Materials for "Linking community water and sanitation access to the global burden of antibiotic resistance using human gut metagenomes from 26 countries"

### Appendix

**List of Tables** **Page**

| List of Tables | Page |
|---|---|
| Table A1: Sources of variables used in this work | 3-4 |
| Table A2: Definitions used for improved and unimproved drinking water and sanitation | 5 |
| Table A3: Study details | 5-6 |
| Table A4: GLM results using 50km and 75km thresholds for household surveys | 8 |
| Table A5: GLM results in the subset of metagenomes with complete metadata | 9 |
| Table A6: GLM results for improved sanitation coverage, improved drinking water coverage, and antibiotic usage | 9 |
| Table A7: GLM results by drug class | 9 |
| Table A8: GLM results in subsets of the data separated by age, World Bank income classification, urbanicity, antibiotic usage in animals, antibiotic usage in humans, relative abundance of *Enterobacteriaceae*, and WHO Region | 10 |
| Table A9: GLM results by individual gene | 10 |
| Table A10: Beta-lactam genes clustered at 80% similarity | 11 |

**List of Figures**

Figure A1: Relative abundance of the five most abundant families of bacteria 6
Figure A2: Total abundance of antibiotic resistance genes by drug class 7
Figure A3: Abundance of antibiotic resistance genes by drug class and study 8

### Methods

Metagenome Identification

The sequence read archives (SRA) was searched for the terms "human gut metagenomes" in the organism field and "metagenomic" in the library source in December 2019 and again in June 2020. Any metagenomes with a geographic location of a low-and middle-income country, according to the World Bank designation in 2021,[1] and short-read sequencing method (Illumina) were retained. A subset of metagenomes from high-income countries were selected to provide diversity in location and age. BioProject and BioSample ID numbers were used to identify corresponding research articles. Results were further refined to include only WGS (whole genome sequencing) assays and exclude amplicon sequencing (e.g., 16S rRNA gene). Longitude and latitude coordinates were obtained from the SRA metagenome metadata and verified with the corresponding research article. If coordinates were unavailable in the SRA, location was determined based on the description in the research article. Additional metadata, when available, were extracted from corresponding research articles including sampling year, age, sex, urban or rural location, sample collection method, DNA extraction method, and library preparation method. The identified metagenome runs were downloaded using SRA Tools (v 2.9.2).

At the time of analysis, additional metagenomes from Mozambique, Kenya, and Bangladesh were contributed by authors from this work. The Mozambique study protocol was approved by the Comité Nacional de Bioética para a Saúde (CNBS), Ministério da Saúde (333/CNBS/14), the Ethics Committee of the London School of Tropical Medicine and Hygiene (reference # 8345), and the Institutional Review Board of the Georgia Institute of Technology (protocol # H15160). The associated MapSan trial has been registered at ClinicalTrials.gov (NCT02362932). The Kenya study protocol was approved by the Committee for the Protection of Human Subjects at the University of California, Berkeley (protocol number 2011-09-3654), the institutional review board at Stanford University (IRB-23310), and the scientific and ethics review unit at the Kenya Medical Research Institute (protocol number SSC-2271). The Bangladesh study protocol for the original trial was approved by the International Centre for Diarrhoeal Diseases Research, Bangladesh (icddr,b) scientific and ethical review committees (protocol number 14022) and the Stanford University human subjects institutional review board (protocol number 30456).

Databases for Antibiotic Resistance Genes and Single Copy Genes

The Comprehensive Antibiotic Resistance Database (CARD)(v 3.0.9)[2] was downloaded in July 2020. While reads were mapped against the full database of homolog models, the CARD entries were clustered at 80% using USEARCH (v 8.1.1861)[3] producing 880 antibiotic resistance gene (ARG) clusters used for naming.

Antibiotic Resistance Gene Identification and Normalization

Downloaded short reads were trimmed using bbmap,[4] including Illumina adapter removal. For paired-end reads, complete pairs were retained. Antibiotic resistance genes were identified by mapping reads to CARD of ARG protein sequences using blastx in DIAMOND (v 0.9.30.131).[5] Results were filtered using a cutoff of 25 amino acids and an identity of 95%. In the case of paired-end reads, forward and reverse reads were mapped separately and results combined, with duplicate mappings removed.

Taxonomy

Kraken2[6] with the standard database (complete genomes in RefSeq for the bacterial, archaeal, and viral domains and Genome Reference Consortium Human Build 38 patch release 13) for taxonomy classification.

Survey Methods

Country-level estimates of GDP were obtained from the World Bank for the year of the study.[7] When the study year was missing, we used data from 2013, as this was the mean year of data collection across all studies. Income classification per country was determined by World Bank designation in 2021.[1] Population density was obtained for the 30 arc-minute region containing the metagenome coordinates from the Center for International Earth Science Information Network at Columbia University.[8] Population density for South Korea was obtained from the World Bank. Estimates for animal antibiotic consumption in 2010 per country were obtained from Our World in Data.[9] Subgroups were determined based on empirical data distributions (antibiotic usage in humans and animals, relative abundance of *Enterobacteriaceae*) or separation by categorical variables that resulted in the most balanced sample sizes (urbanicity, age, WHO region, income classification).

Secondary ARG analysis

To estimate the impact of abundance normalization method on ARG abundance, we conducted a secondary analysis using single copy bacterial genes as an alternative normalization approach. We created a database of genes present in single copy in bacterial genomes (SCGs) by obtaining the sequences of 92 single copy genes from the NCBI's database of clusters of orthologous genes[10] that were present in 95% of bacterial genomes examined in a previous study.[11] Short reads were mapped to the SCG database using BLASTX in DIAMOND with a cutoff of 25 amino acids and an identity of 80%. Paired end reads were mapped separately, and duplicate mappings were removed. Reads per kilobase per million mapped reads (RPKM) of antibiotic resistance genes were normalized by the average RPKM of the 92 SCGs (RPKM ARG/average RPKM of 92 SCGs). Using this approach, the trends in ARG abundance across regions were similar, with the greatest difference in the Americas, where total abundance increased relative to other WHO regions. In the unadjusted model, the association between ARG abundance and improved water and sanitation access was similar and borderline statistically significant accounting for clustered data [-0.10 (-0.26, 0.06), p=0.2].

**Table A1. Sources of variables used in this study.**

| Variable | Source | Level | Units | Countries |
|---|---|---|---|---|
| Access to both Improved Drinking Water and Sanitation | DHS | 25 km radius | % | Bangladesh (2010, 2015), Cameroon (2010, 2015), Egypt (2010), Ethiopia (2010), Ghana (2010), India (2015), Indonesia (2010-province level), Kenya (2015), Liberia (2015), Madagascar (2015), Mozambique (2015), Tanzania (2010, 2015), Peru (2012-province level) |
| | MICS | Province | | El Salvador (2015), Mexico (2015), Mongolia (2015) |
| | Country- specific surveys | Province | | China (2010-Hunan) |
| | Assumed 99% | Country | | Canada, Israel, Italy, USA, Japan, Hong Kong, S. Korea, Netherlands |
| | Study-specific[12] | Study | | Ecuador |
| Toilet Shared | DHS | 25 km radius | % | Bangladesh (2010, 2015), Cameroon (2010, 2015), Egypt (2010), Ethiopia (2010), Ghana (2010), India (2015), Kenya (2015), Liberia (2015), Madagascar (2015), Mozambique (2015), Tanzania (2010, 2015), Peru (2012-province level) |
| | MICS | Province | | El Salvador (2015), Mexico (2015) |
| | Country-specific surveys | Province | | China (2010-Hunan) |
| Antibiotic Consumption in Humans Method 1 | Browne et al.[13] | Country | DDD per 1000 persons per day | Bangladesh, Cameroon, Canada, China, Ecuador, Egypt, El Salvador, India, Indonesia, Italy, Japan, Mexico, Netherlands, Peru, S. Korea, USA, Israel, Ethiopia, Liberia, Ghana, Kenya, Tanzania, Madagascar, Mozambique, Mongolia |
| | Klein et al.[14] | Country | | Hong Kong |
| Antibiotic Consumption in Humans Method 2 (Consumption in children under 5 with illness) | DHS | 25 km radius | % | Bangladesh (2010, 2015), Cameroon (2010, 2015), Egypt (2010), Ethiopia (2010), Ghana (2010), India (2015), |

| | | | | |
|---|---|---|---|---|
| | | | | Indonesia (2010-province level), Kenya (2015), Liberia (2015), Madagascar (2015), Mozambique (2015), Tanzania (2010, 2015), Peru (2012-province level) |
| | MICS | Province | | El Salvador (2015), Mexico (2015), Mongolia (2015) |
| Antibiotic Consumption in Animals | Our world in data[9,15,16] | Country | mg/PCU | Bangladesh, Cameroon, Canada, China, Ecuador, Egypt, Ethiopia, Ghana, India, Indonesia, Israel, Italy, Japan, Kenya, Korea, Liberia, Madagascar, Mexico, Mongolia, Mozambique, Netherlands, Peru, Tanzania, USA |
| GDP per Capita | World Bank | Country | Billion USD/ million people | All |
| Income Classification | World Bank[1] | Country | NA | All |
| Population Density | Center for International Earth Science Information Network at Columbia University[8] | 30-arcminute region | People/km^2 | Bangladesh, Cameroon, Canada, China, Ecuador, Egypt, El Salvador, Ethiopia, Ghana, India, Indonesia, Israel, Italy, Japan, Kenya, Liberia, Madagascar, Mexico, Mongolia, Mozambique, Netherlands, Peru, Tanzania, USA, Hong Kong |
| | World Bank | Country | | S. Korea |
| Household Assets (livestock ownership, finished floors, finished walls, finished roof, electricity access, watch, clock, radio, television, mobile phone, refrigerator, bicycle, motorcycle, car/truck, clean fuel) | DHS | 25 km radius | % | Bangladesh (2010, 2015), Cameroon (2010, 2015), Egypt (2010), Ethiopia (2010), Ghana (2010), India (2015), Indonesia (2010-province level), Kenya (2015), Liberia (2015), Madagascar (2015), Mozambique (2015), Tanzania (2010, 2015), Peru (2012-province level) |
| | MICS | Province | | El Salvador (2015), Mexico (2015) |
| | Country-specific surveys | Province | | China (2010-Hunan) |
| Walk time to drinking water source | DHS | 25 km radius | Minutes | Bangladesh (2010, 2015), Cameroon (2010, 2015), Egypt (2010), Ethiopia (2010), Ghana (2010), India (2015), Indonesia (2010-province level), Kenya (2015), Liberia (2015), Madagascar (2015), Mozambique (2015), Tanzania (2010, 2015), Peru (2012-province level) |
| | MICS | Province | | El Salvador (2015), Mexico (2015) |
| | Country-level | Province | | China (2010-Hunan) |
| Study characteristics (host age, sex, library layout, average read length, rural vs. urban, GPS coordinates) | Sequence read archives and corresponding studies | Sample | NA | All |
| Relative abundance of *Enterobacteriaceae* | Kraken/Bracken[6,17] | Sample | % (Total # of reads classified | All |

| | | | |
|---|---|---|---|
| | | | as Enterobacteriaceae/total number of reads classified as bacteria) |

**Table A2. Definitions used for improved, unimproved, and safely managed drinking water and sanitation.**[18]

| | Improved | Unimproved | Safely Managed |
|---|---|---|---|
| Sanitation | Facilities which ensure hygienic separation of human excreta from human contact (e.g., flush/pour flush to piped sewer, septic tank or pit latrine, composting toilet or pit latrine with slab). | Examples: Pit latrines without a slab or platform, hanging latrines, and bucket latrines. | Use of improved facilities that are not shared with other households and where excreta are safely disposed of in situ or removed and treated offsite. |
| Drinking Water | Drinking water source that by the nature of its construction adequately protects the source from outside contamination, in particular with fecal matter (e.g., piped household connections, public taps or standpipes, boreholes or tube wells, protected dug wells, protected springs, rainwater, tanker trucks, and bottled water). | Examples: Unprotected dug wells, unprotected springs, and surface water. | Improved source located on premises, available when needed, and free from microbiological and priority chemical contamination. |

**Table A3. SRA accession numbers, country, and number of metagenomes for studies included in this analysis.**

| | Study Name | SRA Project No | Country | No. Metagenomes | Citation |
|---|---|---|---|---|---|
| 1 | Microbiome and Worm Infection | PRJNA407815 | Indonesia | 10 | 19 |
| | | | Liberia | 4 | |
| 2 | Indian Human Gut Microbiome | PRJNA397112 | India | 110 | 20 |
| 3 | Comparison of distal gut microbiota structure and function in US and Egyptian children | PRJEB8201 | Egypt | 1 | NA |
| 4 | Subsistence strategies and hunter gatherer microbial communities | PRJNA268964 | Peru | 36 | 21 |
| 5 | Antibiotic resistance exchange between microbiota in resource-poor settings in Latin America | PRJNA300541 | El Salvador | 43 | 22 |
| | | | Peru | 45 | |
| 6 | Metagenome sequencing of the Hadza hunter-gatherer gut microbiota | PRJNA278393 | Tanzania | 27 | 23 |
| 7 | Genome diversity of pathogenic Escherichia coli in Ecuador | PRJNA486009 | Ecuador | 77 | 12 |
| 8 | Stool samples metagenomes from rural communities in Madagascar | PRJNA485056 | Madagascar | 112 | 24 |
| 9 | Metagenomic sequencing of stool samples from Ethiopian individuals | PRJNA504891 | Ethiopia | 50 | 24 |
| 10 | Gut Microbial Succession Follows Acute Secretory Diarrhea in Humans | PRJEB9150 | Bangladesh | 19 | 25 |
| 11 | Antibiotic Treatment Leads to Fecal Escherichia coli and Coliphage Expansion in Severely Malnourished Diarrhea Patients | SRP100895 | Bangladesh | 9 | 26 |
| 12 | Metagenomics analysis reveals features unique to Indian distal gut microbiota | PRJNA531203 | India | 30 | 27 |
| 13 | Gut metagenomes of rural populations in Cameroon | PRJEB27005 | Cameroon | 57 | 28 |
| 14 | Stool samples of a cohort of individuals from Tanzania (Korogwe District) | PRJNA529400 | Tanzania | 68 | 29 |

| | | | | | |
|---|---|---|---|---|---|
| 15 | Metagenomic sequencing of stool samples from Ghanaian individuals | PRJNA529124 | Ghana | 44 | [29] |
| 16 | Seasonal Cycling in the Gut Microbiome of the Hadza Hunter-Gatherers of Tanzania | PRJNA392180 | Tanzania | 19 | [30] |
| 17 | Predicting Vibrio cholera infection and disease severity using metagenomics in a prospective cohort study | PRJNA608678 | Bangladesh | 82 | [31] |
| 18 | Lifestyle and the presence of helminths is associated with gut microbiome composition in Cameroonians | PRJNA547591 | Cameroon | 175 | [32] |
| 19 | The gut microbiome of Mexican children affected by obesity | PRJNA385215 | Mexico | 10 | [33] |
| 20 | Mongolian Metagenome | PRJNA328899 | Mongolia | 62 | [34] |
| 21 | Recent urbanization in China is correlated with a Westernized microbiome encoding increased virulence and antibiotic resistance genes | PRJNA349463 | China | 40 | [35] |
| 22 | Metagenomic analysis of fecal microbiome as a tool towards targeted non-invasive biomarkers for colorectal cancer | ERP012177/PRJEB10 878 | Hong Kong SAR | 50 | [36] |
| 23 | The gut microbiome of healthy Japanese and its microbial and functional uniqueness | PRJDB3601 | Japan | 32 | [37] |
| 24 | Subsistence strategies in traditional societies distinguish gut microbiomes | PRJNA268964 | USA | 14 | [21] |
| 25 | KOALA cohort metagenome study | PRJEB26795 | Netherlands | 50 | [38] |
| 26 | Stability of Gut Enterotypes in Korean Monozygotic Twins and Their Association with Biomarkers and Diet | ERP002391 | S. Korea | 10 | [39] |
| 27 | Personalized Nutrition by Prediction of Glycemic Responses | PRJEB11532 | Israel | 50 | [40] |
| 28 | The initial state of the human gut microbiome determines its reshaping by antibiotics | PRJEB8094 | Canada | 24 | [41] |
| 29 | Mother-to-Infant Microbial Transmission from Different Body Sites Shapes the Developing Infant Gut Microbiome | PRJNA352475 | Italy | 27 | [42] |
| 30 | Dhaka, Bangladesh | PRJNA706606 | Bangladesh | 101 | [43] |
| 31 | Maputo, Mozambique | PRJNA747761 | Mozambique | 85 | NA |
| 32 | Rural Kenya | PRJNA768833 | Kenya | 12 | NA |

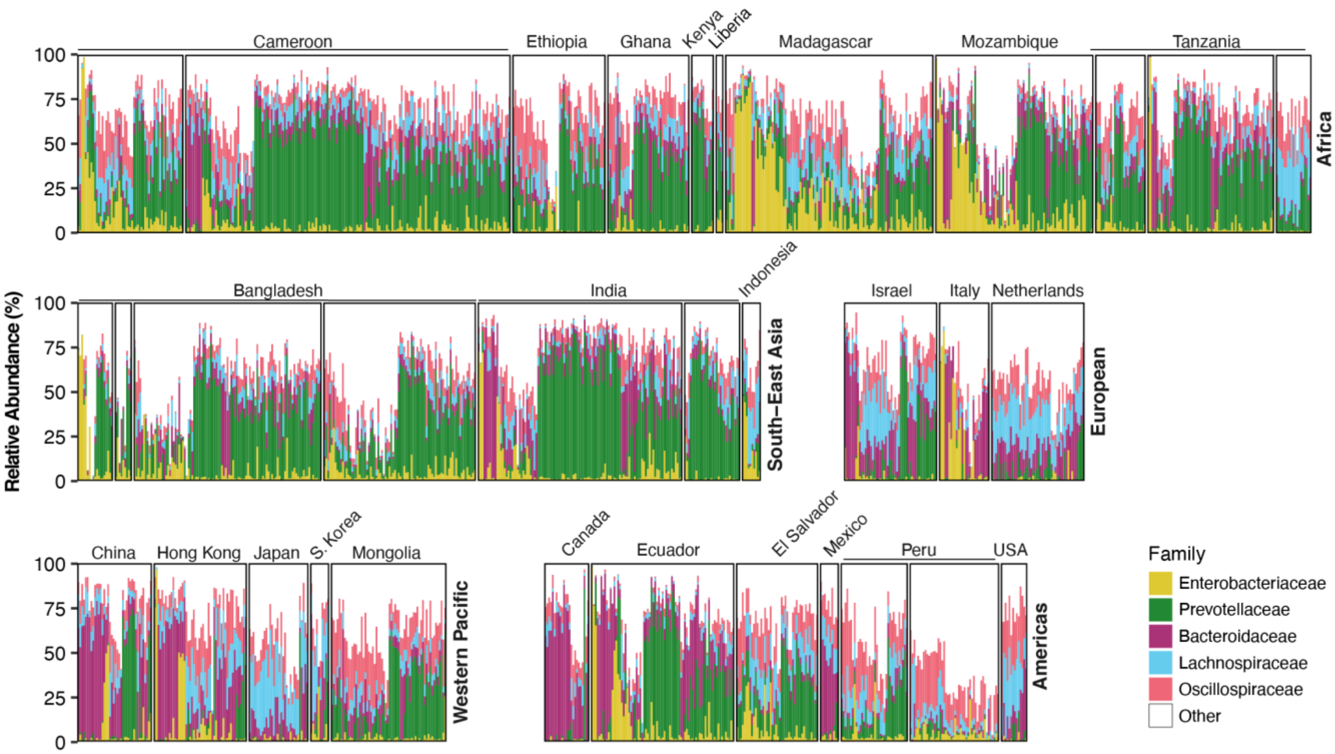

**Figure A1. Relative abundance of the five most abundant families of bacteria normalized by total reads classified as bacteria.** Metagenomes (each bar is one metagenome) ordered by hierarchal clustering of the family level taxa profiles and grouped by study and WHO region. Eastern Mediterranean is not shown due to the presence of only one metagenome (Egypt). Note: multiple studies conducted in Cameroon, Tanzania, Bangladesh, India, and Peru.

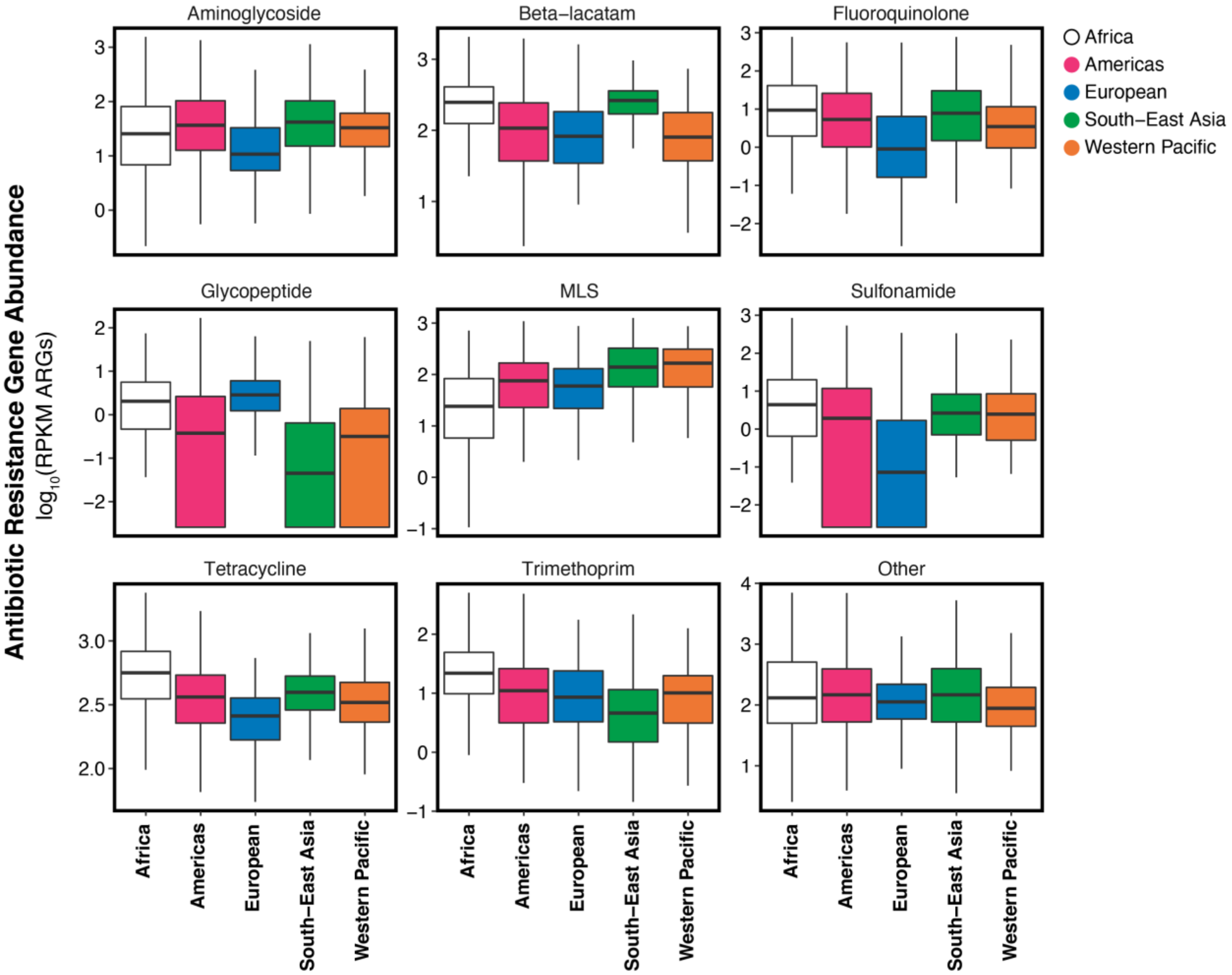

**Figure A2. Total abundance of antibiotic resistance genes in units of $\log_{10}$ (RPKM ARGs using total reads classified as bacteria) by drug class and WHO region.** Boxplots consist of 25$^{th}$ percentile, median, 75$^{th}$ percentile and whiskers extend to at most 1.5x the inner quartile range. Outliers are not shown. Eastern Mediterranean is not shown due to the presence of only 1 metagenome (Egypt). MLS=Macrolide, lincosamide, streptogramin.

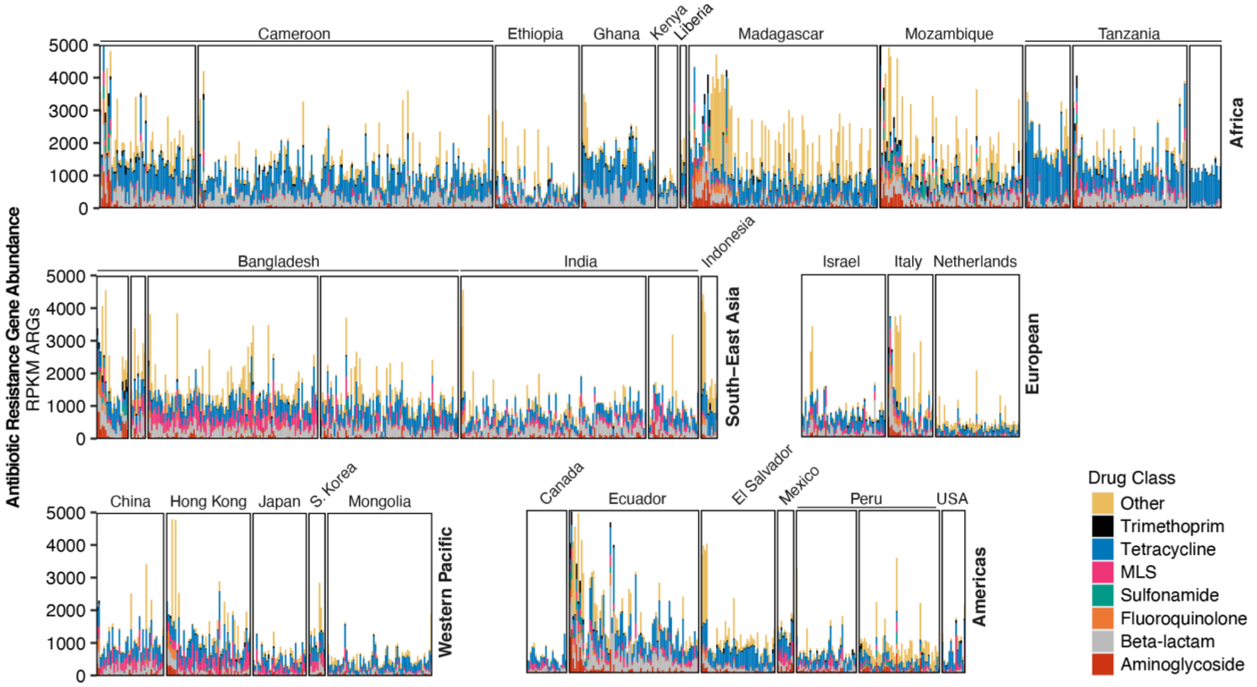

**Figure A3. Abundance of antibiotic resistance genes (RPKM ARGs using total reads classified as bacteria) by drug class and study.** Metagenomes were ordered by hierarchical clustering and grouped by WHO region. 50 metagenomes with abundance > 5000 were removed for visualization. Eastern Mediterranean is not shown due to the presence of only 1 metagenome (Egypt). Note: multiple studies conducted in Cameroon, Tanzania, Bangladesh, India, and Peru.

**Table A4. General linearized model results for total antibiotic resistance gene abundance as a function of combined improved water and sanitation access (improved WASH) using 50km and 75km thresholds for household surveys.**

| | Adjusted All Covariates | | Adjusted WHO Region and Population Density | | |
|---|---|---|---|---|---|
| | **Effect Estimate (95% CI)** | **p-value** | **Effect Estimate (95% CI)** | **p-value** | **N** |
| **50 km** | | | | | |
| | -0.20 (-0.44, 0.03) | 0.09 | -0.21 (-0.43, 0.01) | 0.06 | 1589 |
| 0-25% | Ref | | | | |
| 25-50% | 0.01 (-0.14, 0.16) | 0.89 | -0.02 (-0.16, 0.12) | 0.74 | |
| 50-75% | -0.09 (-0.24, 0.06) | 0.25 | -0.13 (-0.26, 0.01) | 0.06 | |
| 75-100% | -0.17 (-0.35, 0.00) | 0.07 | -0.19 (-0.36, -0.01) | 0.04 | |
| **75 km** | | | | | |
| | -0.22 (-0.48, 0.04) | 0.10 | -0.21 (-0.45, 0.02) | 0.08 | 1589 |
| 0-25% | Ref | | | | |
| 25-50% | 0.02 (-0.14, 0.17) | 0.84 | -0.02 (-0.16, 0.13) | 0.83 | |
| 50-75% | -0.09 (-0.24, 0.07) | 0.26 | -0.12 (-0.26, 0.02) | 0.09 | |
| 75-100% | -0.16 (-0.36, 0.04) | 0.11 | -0.18 (-0.36, 0.01) | 0.06 | |

**Table A5. General linearized model results for total antibiotic resistance gene abundance as a function of combined improved water and sanitation access in the subset of metagenomes with complete metadata.**

| | Adjusted[1] | | Unadjusted | | |
|---|---|---|---|---|---|
| | **Estimate (95% CI)** | **p-value** | **Estimate (95% CI)** | **p-value** | **N** |
| 25 km | -0.39 (-0.65, -0.13) | <0.01 | -0.18 (-0.38, 0.03) | 0.09 | 961 |
| 50 km | -0.18 (-0.76, 0.40) | 0.54 | -0.11 (-0.38, 0.16) | 0.42 | 961 |
| 75 km | -0.34 (-0.75, 0.06) | 0.10 | -0.10 (-0.38, 0.18) | 0.48 | 961 |

**Table A6. General linearized model results for total antibiotic resistance gene abundance as a function of improved sanitation coverage, improved drinking water coverage, and antibiotic usage in humans.** Models were conducted individually. N=1589

| | Adjusted All Covariates | | Adjusted WHO Region | |
|---|---|---|---|---|
| **Independent Variable** | **Effect Estimate (95% CI)** | **p-value** | **Effect Estimate (95% CI)** | **p-value** |
| Sanitation only | -0.16 (-0.32, 0.00) | 0.05 | -0.17 (-0.33, 0.00) | 0.05 |
| Drinking water only | -0.09 (-0.35, 0.16) | 0.47 | -0.10 (-0.29, 0.10) | 0.35 |
| Antibiotic usage only | 0.00 (0.00, 0.01) | 0.34 | 0.00 (-0.01, 0.01) | 0.60 |

**Table A7. General linearized model results for total antibiotic resistance gene abundance in each drug class as a function of combined improved drinking water and sanitation.** Adjusted for WHO region and population density. Drug classes with ≥ 20% of samples considered non-detects are not shown. N=1589

| Drug Class | Estimate (95% CI LB, UB) | p-value |
|---|---|---|
| Tetracycline | -0.31 (-0.48, -0.13) | <0.001 |
| Beta-lactam | -0.27 (-0.64, 0.10) | 0.15 |
| Aminoglycoside | -0.07 (-0.42, 0.28) | 0.70 |
| MLS | 0.08 (-0.41, 0.57) | 0.74 |
| Fluoroquinolone | -0.74 (-1.40, -0.08) | 0.03 |
| Trimethoprim | -1.00 (-1.70, -0.31) | <0.01 |

**Table A8. Association between ARG abundance and combined access to both improved drinking water and sanitation in subsets of the data separated by age, World Bank income classification, urbanicity, antibiotic usage in animals, antibiotic usage in humans, relative abundance of *Enterobacteriaceae*, and WHO Region.** WHO region included as a covariate in all models, except for the WHO region model. Population density included as a covariate for all models, except for urbanicity. Robust standard errors were used to account for clustering (multiple metagenomes at one georeferenced cluster). Interaction p-value for WHO Region determined using a Wald test. PCU: Population-corrected unit. DDD: Defined daily dose. RA: Relative abundance.

| Subgroup | n | Estimate (95% CI) | p-value | Interaction p-value |
|---|---|---|---|---|
| **Age** | | | | |
| 18+ years | 992 | -0.18 (-0.32, -0.05) | 0.01 | 0.71 |
| 0-18 years | 555 | -0.07 (-0.45, 0.32) | 0.74 | |
| **Income** | | | | |
| Low, lower-middle | 1048 | -0.22 (-0.40, -0.05) | 0.01 | 0.60 |
| Upper-middle, high | 546 | -0.31 (-0.66, 0.04) | 0.09 | |
| **Rural vs Urban** | | | | |
| Rural | 825 | -0.16 (-0.38, 0.07) | 0.17 | 0.26 |
| Urban | 704 | -0.37 (-0.68, -0.07) | 0.02 | |
| **Antibiotic Usage in Animals** | | | | |
| < 50 mg/PCU | 834 | -0.22 (-0.42, -0.02) | 0.03 | 0.95 |
| ≥ 50 mg/PCU | 651 | -0.32 (-0.70, 0.07) | 0.10 | |
| **Antibiotic Usage in Humans** | | | | |
| < 12 DDD per 1000 persons per day | 786 | -0.09 (-0.39, 0.21) | 0.55 | 0.68 |
| ≥ 12 DDD per 1000 persons per day | 803 | -0.33 (-0.53, -0.13) | 0.001 | |
| **Relative Abundance of *Enterobacteriaceae*** | | | | |
| RA *Enterobacteriaceae* < 2% | 807 | -0.18 (-0.33, -0.03) | 0.02 | 0.80 |
| RA *Enterobacteriaceae* ≥ 2% | 782 | -0.29 (-0.45, -0.12) | <0.001 | |
| **WHO Region** | | | | |
| Africa | 653 | -0.14 (-0.50, 0.23) | 0.47 | 0.13 |
| Americas | 253 | -0.56 (-0.91, -0.21) | 0.001 | |
| Southeast Asia | 361 | -0.28 (-0.52, -0.03) | 0.03 | |
| Western Pacific | 194 | -0.04 (-0.38, 0.29) | 0.80 | |

**Table A9. Adjusted and unadjusted general linearized model results for abundance of individual antibiotic resistance genes present in at least 5% of metagenomes and significant at p-value <0.05 in the adjusted model.**

| | | Adjusted | | Unadjusted | |
|---|---|---|---|---|---|
| **Gene** | **Drug Class** | **Estimate (95% CI)** | **p-value** | **Estimate (95% CI)** | **p-value** |
| *tetB(P)* | tetracycline | -1.97 (-2.49, -1.45) | <0.001 | -2.00 (-2.70, -1.30) | <0.001 |
| *tetM* | tetracycline | -2.27 (-2.96, -1.58) | <0.001 | -1.97 (-2.66, -1.27) | <0.001 |
| *tetA(P)* | tetracycline | -1.59 (-2.11, -1.08) | <0.001 | -1.53 (-2.26, -0.80) | <0.01 |
| *catS* | phenicol | -1.59 (-2.12, -1.06) | <0.001 | -0.88 (-1.51, -0.25) | 1.00 |
| *C. perfringens mprF* | peptide | -0.46 (-0.63, -0.29) | <0.001 | -0.53 (-0.78, -0.28) | <0.01 |
| OXA Group 12 | beta-lactam | -0.74 (-1.02, -0.46) | <0.001 | -0.56 (-0.86, -0.27) | 0.03 |
| *tetB(60)* | tetracycline | -0.41 (-0.58, -0.25) | <0.001 | -0.20 (-0.41, 0.02) | 1.00 |
| *tet37* | tetracycline | -2.24 (-3.14, -1.34) | <0.001 | -1.94 (-2.92, -0.96) | 0.02 |
| *cmeB* | multi drug | -0.51 (-0.73, -0.30) | <0.001 | -0.50 (-0.71, -0.29) | <0.001 |
| *tetA(60)* | tetracycline | -0.45 (-0.64, -0.26) | <0.001 | -0.24 (-0.46, -0.03) | 1.00 |
| *qnrS* | fluoroquinolone | -1.28 (-1.84, -0.71) | <0.01 | -1.19 (-1.71, -0.68) | <0.01 |
| *tet44* | tetracycline | -1.38 (-2.00, -0.76) | <0.01 | -0.70 (-1.45, 0.06) | 1.00 |
| *ermQ* | MLS | -1.35 (-1.96, -0.74) | <0.01 | -1.09 (-1.70, -0.49) | 0.08 |
| *mtrD* | multi drug | -0.28 (-0.40, -0.15) | <0.01 | -0.27 (-0.39, -0.15) | <0.01 |
| *tetL* | tetracycline | -1.59 (-2.32, -0.86) | <0.01 | -1.20 (-1.91, -0.48) | 0.20 |
| *vanG* | glycopeptide | -1.18 (-1.73, -0.63) | <0.01 | -0.68 (-1.33, -0.03) | 1.00 |
| DHA | beta-lactam | -0.69 (-1.02, -0.37) | <0.01 | -0.65 (-0.95, -0.34) | <0.01 |
| CTX-M Group 2 | beta-lactam | -0.73 (-1.08, -0.38) | <0.01 | -0.67 (-1.05, -0.30) | 0.09 |
| *patB* | fluoroquinolone | -1.06 (-1.57, -0.56) | <0.01 | -0.81 (-1.31, -0.32) | 0.24 |
| *ermT* | MLS | -1.47 (-2.18, -0.77) | <0.01 | -1.23 (-1.95, -0.52) | 0.15 |
| *pmrA* | fluoroquinolone | -0.82 (-1.23, -0.41) | 0.02 | -0.62 (-0.98, -0.25) | 0.19 |
| FosA2 | fosfomycin | -0.56 (-0.84, -0.28) | 0.02 | -0.55 (-0.81, -0.29) | <0.01 |
| RlmA(II) | MLS | -0.78 (-1.17, -0.39) | 0.02 | -0.64 (-0.99, -0.29) | 0.06 |
| *vanXYG* | glycopeptide | -1.17 (-1.77, -0.57) | 0.03 | -0.69 (-1.38, 0.00) | 1.00 |
| *aadA* | aminoglycoside | -1.09 (-1.66, -0.51) | 0.04 | -1.04 (-1.60, -0.48) | 0.05 |
| *msrE* | multi drug | -0.45 (-0.69, -0.21) | 0.05 | -0.44 (-0.71, -0.17) | 0.24 |

**Table A10. Genes clustered at 80% similarity for naming purposes in our analysis. Embolden genes are listed as "current threats" in Zhang et al.[44]**

| Group Name | Genes Clustered into Group |
|---|---|
| OXA Group 12 | OXA-472, OXA-470, OXA-474, OXA-63, OXA-476, OXA-471, OXA-479, OXA-137, OXA-192, OXA-473, OXA-136, OXA-478, OXA-477, OXA-475 |
| CTX-M Group 2 | CTX-M-11, CTX-M-42, CTX-M-144, CTX-M-34, CTX-M-88, CTX-M-116, CTX-M-12, CTX-M-69, CTX-M-114, **CTX-M-15,** CTX-M-136, CTX-M-33, CTX-M-60, CTX-M-139, CTX-M-10, CTX-M-22, CTX-M-82, CTX-M-158, CTX-M-117, CTX-M-62, CTX-M-58, CTX-M-36, CTX-M-32, CTX-M-157, CTX-M-132, CTX-M-109, CTX-M-23, CTX-M-142, CTX-M-68, CTX-M-3, CTX-M-96, **CTX-M-55**, CTX-M-61, CTX-M-54, CTX-M-53, CTX-M-107, CTX-M-108, CTX-M-1, CTX-M-103, CTX-M-30, CTX-M-66, CTX-M-72, CTX-M-52, CTX-M-80, CTX-M-29, CTX-M-156, CTX-M-71, CTX-M-155, CTX-M-123, CTX-M-79, CTX-M-28, CTX-M-37, CTX-M-101 |

**References**

1 World Bank Country and Lending Groups. https://datahelpdesk.worldbank.org/knowledgebase/articles/906519-world-bank-country-and-lending-groups (accessed June 22, 2021).

2 Alcock BP, Raphenya AR, Lau TTY, *et al.* CARD 2020: antibiotic resistome surveillance with the comprehensive antibiotic resistance database. *Nucleic Acids Res* 2020; **48**: D517–25.

3 Edgar RC. Search and clustering orders of magnitude faster than BLAST. *Bioinformatics* 2010; **26**: 2460–1.

4 Bushnell B. BBTools. 2014. https://sourceforge.net/projects/bbmap/.

5 Buchfink B, Xie C, Huson DH. Fast and sensitive protein alignment using DIAMOND. *Nat Methods* 2015; **12**: 59–60.

6 Wood DE, Lu J, Langmead B. Improved metagenomic analysis with Kraken 2. *Genome Biol* 2019; **20**: 257.

7 The World Bank. GDP (current US$) | Data. https://data.worldbank.org/indicator/NY.GDP.MKTP.CD (accessed June 23, 2021).

8 Center For International Earth Science Information Network-CIESIN-Columbia University. Gridded Population of the World, Version 4 (GPWv4): Population Density, Revision 11. 2017. DOI:10.7927/H49C6VHW.

9 Antibiotic use in livestock. Our World Data. https://ourworldindata.org/grapher/antibiotic-use-in-livestock (accessed June 17, 2021).

10 Tatusov RL, Koonin EV, Lipman DJ. A genomic perspective on protein families. *Science* 1997; **278**: 631–7.

11 Na S-I, Kim YO, Yoon S-H, Ha S-M, Baek I, Chun J. UBCG: Up-to-date bacterial core gene set and pipeline for phylogenomic tree reconstruction. *J Microbiol* 2018; **56**: 280–5.

12 Peña-Gonzalez A, Soto-Girón MJ, Smith S, *et al.* Metagenomic Signatures of Gut Infections Caused by Different Escherichia coli Pathotypes. *Appl Environ Microbiol*; **85**: e01820-19.

13 Browne AJ, Chipeta MG, Haines-Woodhouse G, *et al.* Global antibiotic consumption and usage in humans, 2000–18: a spatial modelling study. *Lancet Planet Health* 2021; **5**: e893–904.

14 Klein EY, Van Boeckel TP, Martinez EM, *et al.* Global increase and geographic convergence in antibiotic consumption between 2000 and 2015. *Proc Natl Acad Sci* 2018; **115**: E3463–70.

15 Van Boeckel TP, Brower C, Gilbert M, *et al.* Global trends in antimicrobial use in food animals. *Proc Natl Acad Sci* 2015; **112**: 5649–54.

16 Veterminary Medicines Division. Sales of veterinary antimicrobial agents in 30 European countries in 2015. European Surveillance of Veterinary Antimicrobial Consumption, 2017 https://www.ema.europa.eu/en/documents/report/seventh-esvac-report-sales-veterinary-antimicrobial-agents-30-european-countries-2015_en.pdf.

17 Lu J, Breitwieser FP, Thielen P, Salzberg SL. Bracken: estimating species abundance in metagenomics data. *PeerJ Comput Sci* 2017; **3**: e104.

18 WHO and UNICEF. Progress on drinking-water, sanitation and hygiene: 2017 update and SDG baselines. 2017 https://www.who.int/publications-detail-redirect/9789241512893 (accessed June 23, 2021).

19 Rosa BA, Supali T, Gankpala L, *et al.* Differential human gut microbiome assemblages during soil-transmitted helminth infections in Indonesia and Liberia. *Microbiome* 2018; **6**: 33.

20 Dhakan DB, Maji A, Sharma AK, *et al.* The unique composition of Indian gut microbiome, gene catalogue, and associated fecal metabolome deciphered using multi-omics approaches. *GigaScience* 2019; **8**. DOI:10.1093/gigascience/giz004.

21 Obregon-Tito AJ, Tito RY, Metcalf J, *et al.* Subsistence strategies in traditional societies distinguish gut microbiomes. *Nat Commun* 2015; **6**: 6505.

22 Pehrsson EC, Tsukayama P, Patel S, *et al.* Interconnected microbiomes and resistomes in low-income human habitats. *Nature* 2016; **533**: 212–6.

23 Rampelli S, Schnorr SL, Consolandi C, *et al.* Metagenome Sequencing of the Hadza Hunter-Gatherer Gut Microbiota. *Curr Biol* 2015; **25**: 1682–93.

24 Pasolli E, Asnicar F, Manara S, *et al.* Extensive Unexplored Human Microbiome Diversity Revealed by Over 150,000 Genomes from Metagenomes Spanning Age, Geography, and Lifestyle. *Cell* 2019; **176**: 649-662.e20.

25 David LA, Weil A, Ryan ET, *et al.* Gut Microbial Succession Follows Acute Secretory Diarrhea in Humans. *mBio*; **6**: e00381-15.

26 Kieser S, Sarker SA, Berger B, *et al.* Antibiotic Treatment Leads to Fecal Escherichia coli and Coliphage Expansion in Severely Malnourished Diarrhea Patients. *Cell Mol Gastroenterol Hepatol* 2018; **5**: 458-460.e6.

27 Kaur K, Khatri I, Akhtar A, Subramanian S, Ramya TNC. Metagenomics analysis reveals features unique to Indian distal gut microbiota. *PLOS ONE* 2020; **15**: e0231197.

28 Lokmer A, Cian A, Froment A, *et al.* Use of shotgun metagenomics for the identification of protozoa in the gut microbiota of healthy individuals from worldwide populations with various industrialization levels. *PLOS ONE* 2019; **14**: e0211139.

29 Tett A, Huang KD, Asnicar F, *et al.* The Prevotella copri Complex Comprises Four Distinct Clades Underrepresented in Westernized Populations. *Cell Host Microbe* 2019; **26**: 666-679.e7.

30 Smits SA, Leach J, Sonnenburg ED, *et al.* Seasonal cycling in the gut microbiome of the Hadza hunter-gatherers of Tanzania. *Science* 2017; **357**: 802–6.

31 Levade I, Saber MM, Midani FS, *et al.* Predicting Vibrio cholerae Infection and Disease Severity Using Metagenomics in a Prospective Cohort Study. *J Infect Dis* 2021; **223**: 342–51.

32 Rubel MA, Abbas A, Taylor LJ, *et al.* Lifestyle and the presence of helminths is associated with gut microbiome composition in Cameroonians. *Genome Biol* 2020; **21**: 122.

33 Maya-Lucas O, Murugesan S, Nirmalkar K, *et al.* The gut microbiome of Mexican children affected by obesity. *Anaerobe* 2019; **55**: 11–23.

34 Liu W, Zhang J, Wu C, *et al.* Unique Features of Ethnic Mongolian Gut Microbiome revealed by metagenomic analysis. *Sci Rep* 2016; **6**: 34826.

35 Winglee K, Howard AG, Sha W, *et al.* Recent urbanization in China is correlated with a Westernized microbiome encoding increased virulence and antibiotic resistance genes. *Microbiome* 2017; **5**: 121.

36 Yu J, Feng Q, Wong SH, *et al.* Metagenomic analysis of faecal microbiome as a tool towards targeted non-invasive biomarkers for colorectal cancer. *Gut* 2017; **66**: 70–8.

37 Nishijima S, Suda W, Oshima K, *et al.* The gut microbiome of healthy Japanese and its microbial and functional uniqueness. *DNA Res* 2016; **23**: 125–33.

38 Zhong H, Penders J, Shi Z, *et al.* Impact of early events and lifestyle on the gut microbiota and metabolic phenotypes in young school-age children. *Microbiome* 2019; **7**: 2.

39 Lim MY, Rho M, Song Y-M, Lee K, Sung J, Ko G. Stability of Gut Enterotypes in Korean Monozygotic Twins and Their Association with Biomarkers and Diet. *Sci Rep* 2014; **4**: 7348.

40 Zeevi D, Korem T, Zmora N, *et al.* Personalized Nutrition by Prediction of Glycemic Responses. *Cell* 2015; **163**: 1079–94.

41 Raymond F, Ouameur AA, Déraspe M, *et al.* The initial state of the human gut microbiome determines its reshaping by antibiotics. *ISME J* 2016; **10**: 707–20.

42 Ferretti P, Pasolli E, Tett A, *et al.* Mother-to-Infant Microbial Transmission from Different Body Sites Shapes the Developing Infant Gut Microbiome. *Cell Host Microbe* 2018; **24**: 133-145.e5.

43 Nadimpalli ML, Lanza VF, Montealegre MC, *et al.* Drinking water chlorination has minor effects on the intestinal flora and resistomes of Bangladeshi children. *Nat Microbiol* 2022; **7**: 620–9.

44 Zhang A-N, Gaston JM, Dai CL, *et al.* An omics-based framework for assessing the health risk of antimicrobial resistance genes. *Nat Commun* 2021; **12**: 4765.
